# The efficacy of convalescent plasma for the treatment of severe influenza

**DOI:** 10.1101/2020.02.20.20025593

**Authors:** Zhiheng Xu, Jianmeng Zhou, Yongbo Huang, Xuesong Liu, Yonghao Xu, Sibei Chen, Dongdong Liu, Zhimin Lin, Xiaoqing Liu, Yimin Li

## Abstract

**Background:** Administration of convalescent plasma may be of clinical benefit for treatment of severe acute viral respiratory infections. However, no clear evidence exists to support or oppose convalescent plasma use in clinical practice. We conducted a systematic review and meta-analysis to assess the evidence of randomized controlled trials (RCTs) in the convalescent plasma for the treatment of severe influenza.

**Methods:** Healthcare databases were searched in February 2020. All records were screened against the eligibility criteria. Data extraction and risk of bias assessments were undertaken. The primary outcome was case-fatality rates by influenza.

**Results:** We identified 5 RCTs of severe influenza. The pooled analyses showed no evidence for a reduction in mortality (Odds Ratio (OR) 1.06; 95% confidence interval (CI) 0.51–2.23; p = 0.87; I^2^ = 35%). We also found non-significant reductions in days in ICU and hospital, and days on mechanical ventilation. There seemed to have a biological benefit of increasing HAI titer levels and decreasing influenza B virus loads and cytokines after convalescent plasma treatment. No serious adverse events was reported between two groups. Studies were commonly of low risk of bias with high quality.

**Conclusions:** Convalescent plasma appears safe but may not reduce mortality in severe influenza. This therapy should be studied within the context of a well-designed clinical trial for treatment of SARS-Cov-2 infection.

## Introduction

Viral pneumonia is a major cause of morbidity and mortality around the world (1, 2). The causative organisms for viral pneumonia vary greatly. The emergence of severe acute respiratory syndrome coronaviruses (SARS-CoV) (3), avian influenza A (H5N1) virus (4) and the Middle East respiratory syndrome coronaviruses (MERS-CoV) (5) played an important role as the causes of severe pneumonia successively. And now attention is turning to a novel coronavirus (SARS-CoV-2).

Since December 2019, an increasing number of cases of pneumonia infected by SARS-CoV-2 have been reported in China (6-8). Up to our knowledge, there was no effective antiviral for the infection. The current approach to clinical management was general supportive care, provided with critical care and organ support when necessary.

It has been suggested that administration of high-titre anti-influenza immune plasma, derived from convalescent or immunised individuals, may yield a clinical effect for treatment of seasonal and pandemic influenza of viral etiology (9-11). It was reported that convalescent plasma treatment reduced the hospital stay and mortality in patients with SARS-CoV infection and severe influenza A (H1N1) (9, 12). Furthermore, systematic reviews of studies using convalescent plasma also found evidence of clinical benefit for such patients (9, 13). Therefore, convalescent plasma may be promising in patients with SARS-CoV-2 infection (COVID-19), particularly in those presented with critical illness.

However, the underlying evidence based on previous studies was of poor quality because few of them was randomized trial. And recently, randomized controlled trials had shown that convalescent plasma or hyperimmune intravenous immunoglobulin (H-IVIG) prepared from pooled plasma obtained from convalescent patients conferred no significant benefit over placebo for patients with influenza infection (14, 15), which was contrast to the previous studies.

Therefore, we conducted a systematic review and meta-analysis to evaluate the clinical efficacy of either convalescent plasma or hyperimmune immunoglobulin for the treatment of severe influenza, to help inform clinical management of SARS-CoV-2 infection.

## METHODS

### Inclusion and exclusion criteria

We included prospective randomized controlled trials (RCTs) involving patients with influenza treated by convalescent plasma and hyperimmune immunoglobulin. We limited publications to the English language. We excluded crossover trials, before-after studies, conference presentations, abstract publications, case reports and editorials. Case series or other studies with no comparator will also be excluded.

### Search strategy

Two reviewers (Z.H.X and J.M.Z) executed the search strategy in February 2020. To increase the sensitivity of our search strategy, we combined the terms “influenza” with “convalescent plasma” or “convalescent serum” or “hyperimmune immunoglobulin” or “immune plasma” or “H-IVIG” as key words or Medical Subject Headings (MeSH) terms. We searched 4 databases (Pubmed, EMBASE, Scopus, and Web of Science) from electronic databases inception to February 10th, 2020. We systematically screened abstracts and full text publications for studies that met our eligibility criteria.

### Definitions

The study population of interest was patients of any age or sex who were hospitalized with influenza with a laboratory-confirmed viral infection. The intervention was convalescent plasma, serum, or hyperimmune immunoglobulin derived from convalescent or immunised individuals. Comparator treatments included placebo, low titre plasma, or sham therapy.

### Outcomes

The primary outcome of this review were case-fatality rates by influenza, reflecting the efficacy of convalescent plasma and hyperimmune immunoglobulin therapy. Secondary outcomes included antibody levels, cytokine levels, viral loads, incidence of serious adverse events, days on mechanical ventilation, ICU and hospital.

### Data abstraction

Two investigators (Z.H.X and J.M.Z) independently reviewed and abstracted data from each retrieved article and supplement. Discrepancies were resolved by discussion and consensus.

### Quality assessment

We assessed the quality of all included trials based on review of the details in the method section and supplements of included trials. We appraised trial quality using the Cochrane collaboration tool for assessing risk of bias (RoB) (16) including assessment of random sequence generation, allocation concealment, blinding (of interventions and outcome measurement or assessment), incomplete outcome data, selective reporting bias and other potential sources of bias (e.g., industry funding). For each criterion, we appraised the RoB to be either of low, high, or unclear risk (e.g., insufficient details). Two authors (Z.H.X, J.Z.M) independently assessed study quality and disagreements were resolved by consensus.

### Assessment of heterogeneity

We used the I^2^ statistic to evaluate the impact of heterogeneity on pooled results. An I^2^ value of greater than 50% indicated substantial heterogeneity (16). We used fixed-effects models to pool data when heterogeneity was insignificant and the random effects models to pool data when significant heterogeneity was identified.

### Statistical analysis

Categorical data was pooled using the odds ratios (ORs), with the 95% confidence intervals (CIs). Statistical analyses were conducted with Review Manager (RevMan) Version 5.3 (Copenhagen: The Nordic Cochrane Centre, The Cochrane Collaboration, 2014), and two-sided p values < 0.05 were considered statistically significant.

## Results

### Description of studies

We identified 2,861 potentially eligible studies. After exclusion of duplicate and irrelevant articles, 29 trials were retrieved to be reviewed in greater detail. Of these, we excluded 24 studies that did not meet our eligibility criteria and thus included 5 trials in our review (Fig.1).

**Figure 1.**
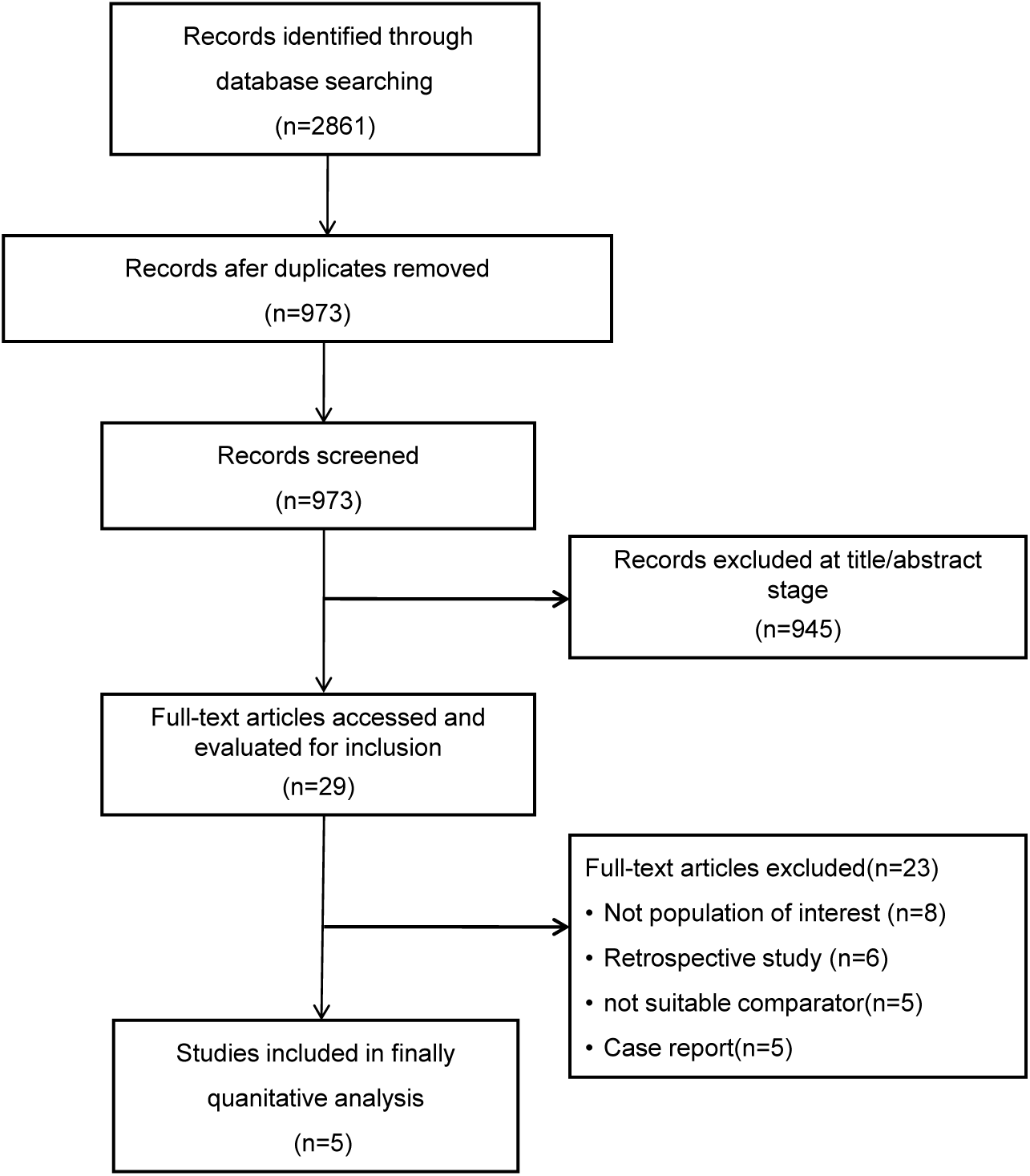
Search strategy of meta-analysis on selecting patients for inclusion.

All of the 5 studies were multicenter randomized controlled trials. Hung et al had found that hyperimmune IV immunoglobulin (H-IVIG) administrated within 5 days of symptom onset was associated with a lower viral load and reduced mortality in patients with severe H1N1 infection (17). The pilot study from INSIGHT FLU005 IVIG Pilot Study Group showed that H-IVIG administration significantly increased hemagglutination inhibition (HAI) titer levels among patients with influenza (18). Later, they performed an international, double-blind RCT, which shown that hIVIG had similar safety outcome of death and adverse events (15). Furthermore, Beigel et al did a multicenter phase 2 trial and found that immune plasma provided support for a possible benefit of severe influenza (19). Recently, their phase 3 trial had shown that high-titre anti-influenza plasma conferred no significant benefit of severe influenza A (14) (Table 1).

**Table 1.**
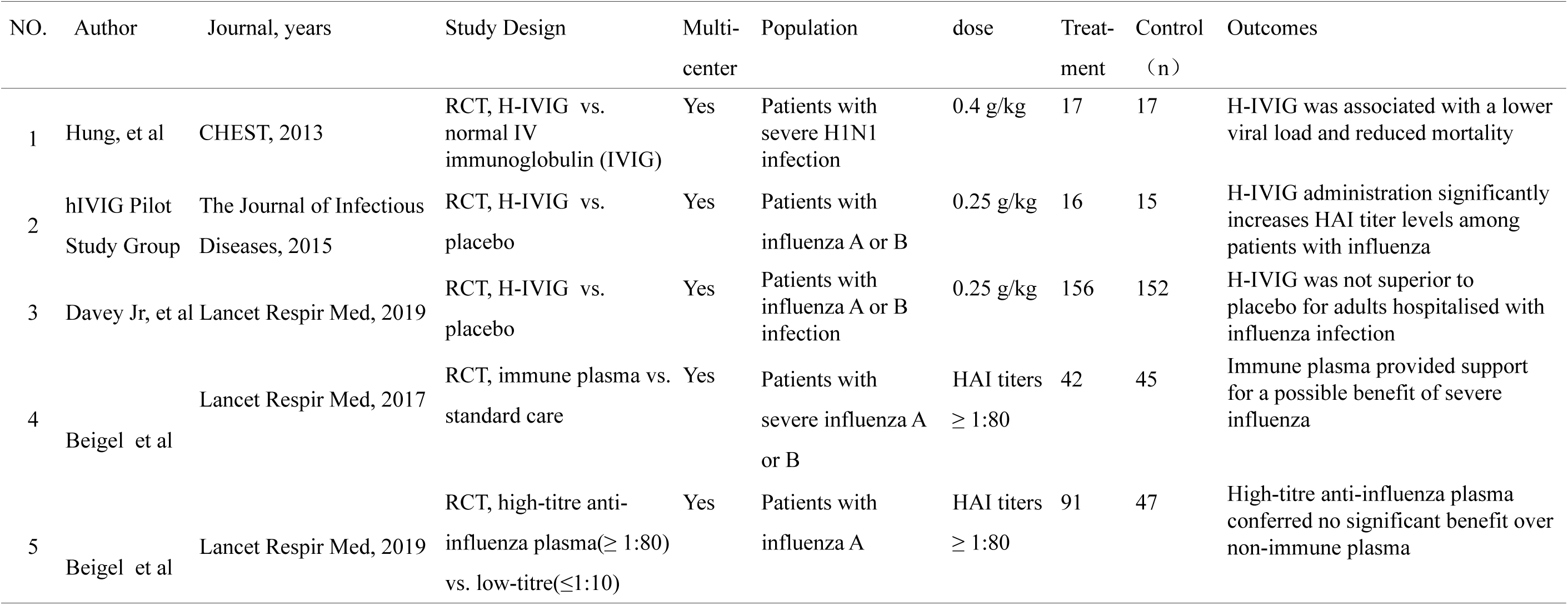
Characteristics of included studies

### Risk of bias of included studies

The RCTs included were all assessed to be at low risk of bias with respect to attribution bias, reporting bias and selection bias except for 1 trial for which random sequence generation was deemed unclear (18). The phase 2 trial of Beigel et al was an open-label study, which meant no binding was performed and resulting high risk of allocation concealment, performance bias and detection bias (19). However, the phase 3 trial of Beigel et al was a multi-center, randomised, double-blind study with low risk of attribution, reporting and selection bias (14). The trial of Davey Jr et al was found to be at low risks in dealing with every aspect (15) (Fig.2).

**Figure 2.**
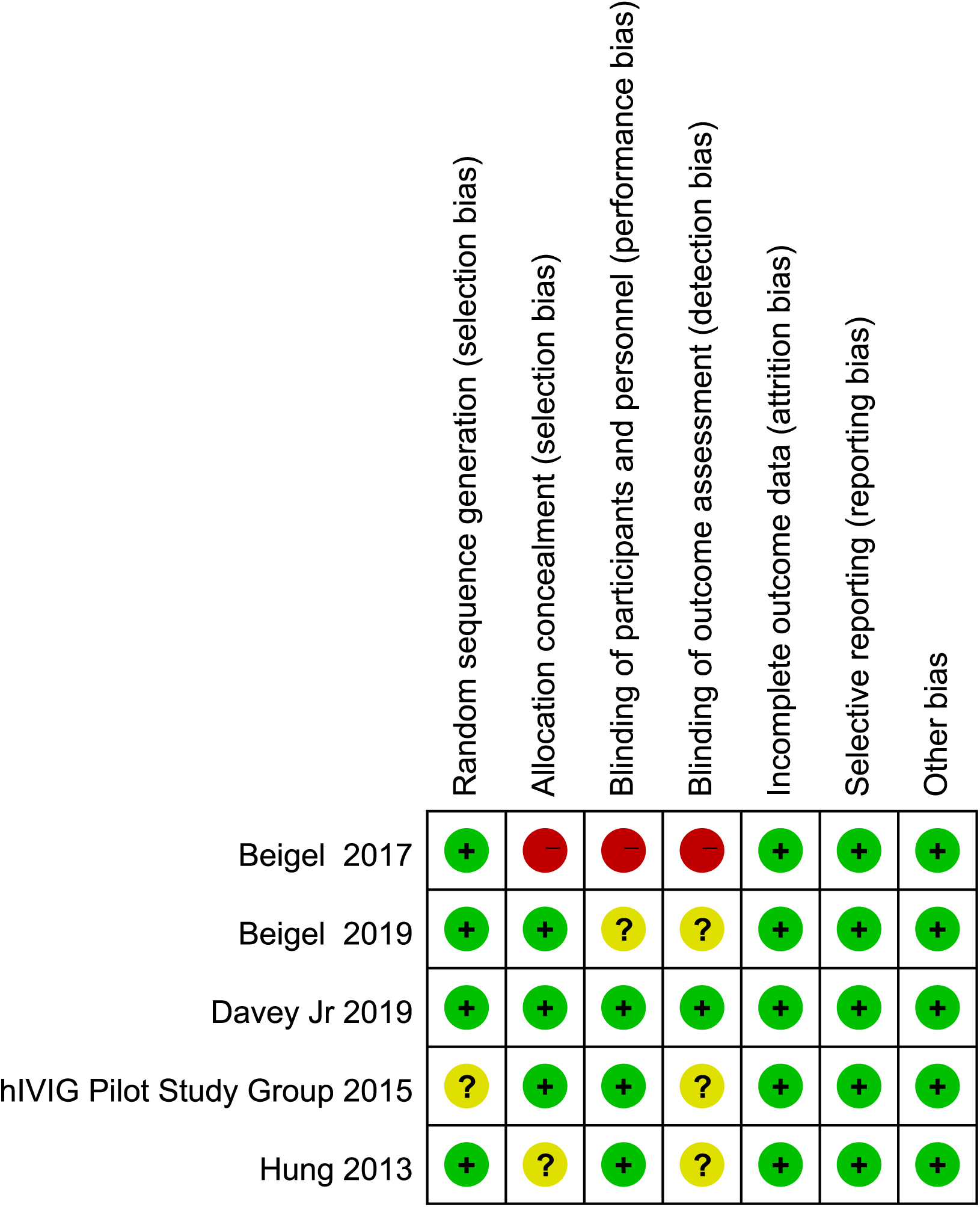
Risk of bias diagram for each study. Green represents low risk of bias, yellow represents unclear risk of bias, and red represents high risk of bias.

### Primary outcome of mortality

There were 4 trials with extractable data included to assess the efficacy in reducing mortality of severe influenza by immune plasma (14, 15, 17, 19). The pooled data (n=567) showed that immune plasma/H-IVIG had no significant difference in reducing the rate of deaths compared with placebo in patients with severe influenza (OR 1.06; 95%CI 0.51–2.23; p = 0.87; I^2^ = 35%) (Fig.3).

**Figure 3.**
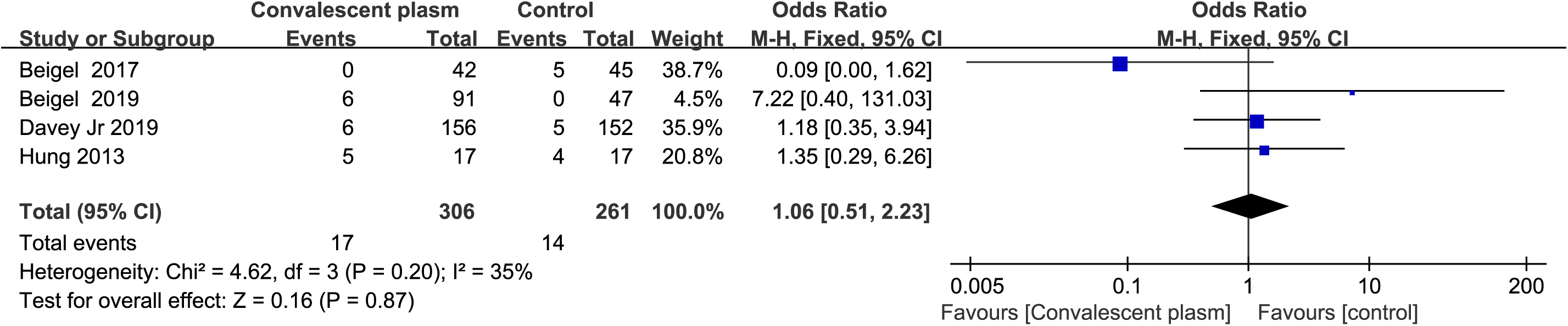
Pooled estimates of case-fatality rates of convalescent plasma compared with control in patients with severe influenza.

## Secondary outcomes

### Antibody Levels

It was reported that HAI titer levels significantly increased in the patients infected with influenza A and influenza B receiving H-IVIG. However, HAI titer levels decreased gradually after the first week treatment (15, 18) (Table 2).

**Table 2.** Secondary outcomes

| Secondary outcomes | Author | hIVIG/immune plasma group | Control group | P value |
| --- | --- | --- | --- | --- |
| Antibody levels | Davey Jr, et al, 2019;<br>hIVIG Pilot Study Group, 2016 | significantly increases HAI titer levels among patients with influenza A and B | -- | -- |
| Viral loads | Hung et al, 2013 | 3.3 log 10 copies/mL(H1N1) | 4.67 log 10 copies/mL | 0.04 |
|  | Davey Jr, et al, 2019 | Mean log10 RNA -1.95(Influenza A) | -2.62 | 0.02 |
|  | Davey Jr, et al, 2019 | Mean log10 RNA -2.09(Influenza B) | -1.54 | 0.005 |
|  | Beigel et al, 2017 | Median log10 copies per mL 1.9 (1.9–1.9) day7<br>( Nasal swab, Influenza A and B ) | 1.9 (1.9–1.9) | NS |
| Cytokines | Hung et al, 2013 | TNF-a, IL-1ra and IL-10 fell to a similar level as control 3 days after treatment | -- | -- |
| Days on mechanical ventilation | Beigel et al, 2017 | 0 (0–6) (Influenza A and B) | 3 (0–14) | 0.14 |
|  | Beigel et al, 2019 | 9 (4–16) (Influenza A) | 15.5 (7.0–29.0) | 0.22 |
| Length of ICU stay, day | Hung et al, 2013 | 11 (4-13.5)(H1N1) | 10 (4.5-13.5) | NS |
|  | Beigel et al, 2017 | 2.5 (0.0–9.0) (Influenza A and B) | 3 (0–13) | 0.37 |
|  | Beigel et al, 2019 | 5.0(3.0–12.5) (Influenza A) | 8 (4–25) | 0.32 |
| Length of hospital stay, day | Hung et al, 2013 | 16 (11.5-13.5)(H1N1) | 16 (7-29) | NS |
|  | Beigel et al, 2017 | 6 (4–16) (Influenza A and B) | 11 (5–25) | 0.13 |
|  | Beigel et al, 2019 | 5 (3–12) (Influenza A) | 6 (4–12) | 0.30 |
| Serious adverse events | Beigel et al, 2017 | 20%(Influenza A and B) | 38% | 0.041 |
|  | Beigel et al, 2019 | 35% (Influenza A ) | 32% | NS |

### Viral Loads and Cytokines

Hung et al had found that H1N1 viral loads were significantly lower in patients after convalescent plasma infused than in the control group. Furthermore, the cytokines, including IL-1ra, IL-10, and TNF-a, were also lower in the convalescent plasma treatment group (17). However, in another large clinical trial, overall viral loads were decreased in both H-IVIG and placebo group during the first 3 days (p=0.49). There was 16% of H-IVIG group and 20% of placebo group had no detectable virus (p=0.15) after infusion. For subgroup with influenza B, the decline in viral loads was greater for the H-IVIG group than for the placebo group (p=0·053) (15). In addition, there was no significant difference in influenza A and B viral loads and the time to virus becoming undetectable (19) (Table 2).

### Length of ICU/ Hospital Stay

Trials from Hung et al and Beigel et al had found that there was no significant difference in the length of ICU stay between the H-IVIG/immune plasma treatment group and the control group. Moreover, the length of hospital stay was similar between the two groups (14, 17, 19). In addition, days on mechanical ventilation was similar between immune plasma treatment and standard care alone (14, 19). Furthermore, there was no different in the proportion of patients alive and discharged at day 7 and day 28 in the two groups (15) (Table 2).

### Serious Adverse Events

No adverse events related to treatment were reported in Hung’s trial (17). In addition, compared to placebo, H-IVIG didn’t show serious adverse events in the FLU-IVIG Trial (15). Fewer participants with immune plasma infusion had serious adverse events in an open-label RCT (19), but later double-blind trial showed similar serious adverse events in the two groups, the most frequent of which were acute respiratory distress syndrome (14, 19) (Table 2).

## Discussion

Our analyses suggested that convalescent plasma may not have a clinically relevant impact in reducing the rate of mortality in patients with influenza. We also found non-significant reductions in days in ICU and hospital, and days on mechanical ventilation. Of interest was the evidence for a benefit of increasing HAI titer levels and decreasing influenza B virus loads and cytokines after convalescent plasma treatment. No serious adverse events was reported.

The use of immune plasma has been recommended as a primary therapy for severe respiratory infectious diseases, including influenza, SARS, and MERS (9, 13, 19). However, data supporting these recommendations are weak and limited to case reports, case series without control. For patients with Coronavirus Disease 2019 (COVID-19), we did not identify any report of convalescent plasma use for these patients in publication. Up to present, there an open-label, non-randomized clinical trial (NCT04264858) and a multicenter, randomized and parallel controlled trial (ChiCTR2000029757) was designed and registered. However, it may take time to perform such clinical trials in recruiting patients and collecting convalescent plasma. Therefore, in order to provide implications of convalescent plasma for COVID-19 clinical practice, we try to carry out a meta-analysis and systemic review only included RCTs of influenza to better evaluate the efficacy of convalescent plasma. We included 5 high quality RCTs of convalescent plasma and H-IVIG use in severe influenza. The evidence for a reduction in mortality associated with convalescent plasma was strongest for influenza A (H1N1) (17). But we should interpret this with an appropriate level of caution because of the limited sample size (n=17) and early use (onset within 5 days). In addition, the pooled data of 4 trails (n=567) that reporting death showed no benefit for reducing the mortality of severe influenza with the treatment.

For secondary outcomes, including days in ICU and hospital, and days on mechanical ventilation, 3 RCTs reported relevant data showed non-significant reductions between H-IVIG/immune plasma group and control group (14, 17, 19). Despite robust increases in HAI titres for influenza A and B (15, 18), decreases influenza B viral loads (15) and cytokines levels in H1N1 (17), there was no clinical benefit observed in patients receiving H-IVIG/immune plasma infusion.

According to the findings of high quality RCTs, we did not identify passive immunotherapy as an adjunctive therapy providing clinical benefit for patients with severe influenza. Therefore, for patients with COVID-19, clinicians should take such previous findings carefully into considerations before the use of convalescent plasma in critically ill SARS-CoV-2 infected patients. The composition of the plasma is complex. And the antibodies produced by the human body are matched with its own immune system. Transfusion reactions may occur in blood products administration (20, 21). In addition, the titers of the antibody from convalescent may differ from each other. The standardized extraction and purification specific antibody may be a difficult and time consuming work, which might not suitable for the current outbreak of SARS-CoV-2. Finally, pilot study and control trials should be carried out to identify the optimal timing, dosage and indications in the use of H-IVIG/immune plasma in patients with COVID-19.

## Conclusion

Available high quality evidence suggested that convalescent plasma/H-IVIG was safe but unlikely to reduce mortality in patients with severe influenza. Despite the current outbreak and emergency of SARS-CoV-2 affecting public health, convalescent plasma should be carefully taken into consideration before a well-designed clinical trial was carried out.

## Data Availability

The authors declare that all data of this study are available from the included 5 RCTs and its supplementary information files.

## Notes

### Competing Interest Statement

The authors have declared no competing interest.

### Clinical Trial

not application

### Funding Statement

National Science and Technology Major Project (No. 2017ZX10204401); National Natural Science Foundation of China (81970071); the Special Project for Emergency of the Ministry of Science and Technology (2020YFC0841300); the Special Project of Guangdong Science and Technology Department (2020B111105001).

## Reference

1. Ruuskanen O, Lahti E, Jennings LC, Murdoch DR. Viral pneumonia. Lancet 2011; 377: 1264–1275.

2. Dandachi D, Rodriguez-Barradas MC. Viral pneumonia: etiologies and treatment. J Investig Med 2018; 66: 957–965.

3. Drosten C, Günther S, Preiser W, van der Werf S, Brodt H-R, Becker S, Rabenau H, Panning M, Kolesnikova L, Fouchier RAM, Berger A, Burguière A-M, Cinatl J, Eickmann M, Escriou N, Grywna K, Kramme S, Manuguerra J-C, Müller S, Rickerts V, Stürmer M, Vieth S, Klenk H-D, Osterhaus ADME, Schmitz H, Doerr HW. Identification of a novel coronavirus in patients with severe acute respiratory syndrome. The New England journal of medicine 2003; 348: 1967–1976.

4. Lai S, Qin Y, Cowling BJ, Ren X, Wardrop NA, Gilbert M, Tsang TK, Wu P, Feng L, Jiang H, Peng Z, Zheng J, Liao Q, Li S, Horby PW, Farrar JJ, Gao GF, Tatem AJ, Yu H. Global epidemiology of avian influenza A H5N1 virus infection in humans, 1997- 2015: a systematic review of individual case data. Lancet Infect Dis 2016; 16: e108–e118.

5. Zumla A, Hui DS, Perlman S. Middle East respiratory syndrome. Lancet 2015; 386: 995–1007.

6. Hui DS, I Azhar E, Madani TA, Ntoumi F, Kock R, Dar O, Ippolito G, McHugh TD, Memish ZA, Drosten C, Zumla A, Petersen E. The continuing 2019-nCoV epidemic threat of novel coronaviruses to global health - The latest 2019 novel coronavirus outbreak in Wuhan, China. Int J Infect Dis 2020; 91: 264–266.

7. Lu H, Stratton CW, Tang Y-W. Outbreak of Pneumonia of Unknown Etiology in Wuhan China: the Mystery and the Miracle. J Med Virol 2020: 10.1002/jmv.25678.

8. Zhu N, Zhang D, Wang W, Li X, Yang B, Song J, Zhao X, Huang B, Shi W, Lu R, Niu P, Zhan F, Ma X, Wang D, Xu W, Wu G, Gao GF, Tan W, China Novel Coronavirus I, Research T. A Novel Coronavirus from Patients with Pneumonia in China, 2019. The New England journal of medicine 2020: 10.1056/NEJMoa2001017.

9. Mair-Jenkins J, Saavedra-Campos M, Baillie JK, Cleary P, Khaw F-M, Lim WS, Makki S, Rooney KD, Nguyen-Van-Tam JS, Beck CR, Convalescent Plasma Study G. The effectiveness of convalescent plasma and hyperimmune immunoglobulin for the treatment of severe acute respiratory infections of viral etiology: a systematic review and exploratory meta-analysis. J Infect Dis 2015; 211: 80–90.

10. Simmons CP, Bernasconi NL, Suguitan AL, Mills K, Ward JM, Chau NVV, Hien TT, Sallusto F, Ha DQ, Farrar J, de Jong MD, Lanzavecchia A, Subbarao K. Prophylactic and therapeutic efficacy of human monoclonal antibodies against H5N1 influenza. PLoS Med 2007; 4: e178–e178.

11. Wu JT, Lee CK, Cowling BJ, Yuen KY. Logistical feasibility and potential benefits of a population-wide passive-immunotherapy program during an influenza pandemic. Proc Natl Acad Sci U S A 2010; 107: 3269–3274.

12. Hung IF, To KK, Lee C-K, Lee K-L, Chan K, Yan W-W, Liu R, Watt C-L, Chan W-M, Lai K-Y, Koo C-K, Buckley T, Chow F-L, Wong K-K, Chan H-S, Ching C-K, Tang BS, Lau CC, Li IW, Liu S-H, Chan K-H, Lin C-K, Yuen K-Y. Convalescent plasma treatment reduced mortality in patients with severe pandemic influenza A (H1N1) 2009 virus infection. Clin Infect Dis 2011; 52: 447–456.

13. Luke TC, Kilbane EM, Jackson JL, Hoffman SL. Meta-analysis: convalescent blood products for Spanish influenza pneumonia: a future H5N1 treatment? Ann Intern Med 2006; 145: 599–609.

14. Beigel JH, Aga E, Elie-Turenne M-C, Cho J, Tebas P, Clark CL, Metcalf JP, Ozment C, Raviprakash K, Beeler J, Holley HP, Jr., Warner S, Chorley C, Lane HC, Hughes MD, Davey RT, Jr., Team IRCS. Anti-influenza immune plasma for the treatment of patients with severe influenza A: a randomised, double-blind, phase 3 trial. Lancet Respir Med 2019; 7: 941–950.

15. Davey RT, Jr., Fernández-Cruz E, Markowitz N, Pett S, Babiker AG, Wentworth D, Khurana S, Engen N, Gordin F, Jain MK, Kan V, Polizzotto MN, Riska P, Ruxrungtham K, Temesgen Z, Lundgren J, Beigel JH, Lane HC, Neaton JD, Group IF-IS.Anti-influenza hyperimmune intravenous immunoglobulin for adults with influenza A or B infection (FLU-IVIG): a double-blind, randomised, placebo-controlled trial. Lancet Respir Med 2019; 7: 951–963.

16. Higgins JPT, Altman DG, Gøtzsche PC, Jüni P, Moher D, Oxman AD, Savovic J, Schulz KF, Weeks L, Sterne JAC, Cochrane Bias Methods G, Cochrane Statistical Methods G. The Cochrane Collaboration’s tool for assessing risk of bias in randomised trials. BMJ 2011; 343: d5928–d5928.

17. Hung IFN, To KKW, Lee C-K, Lee K-L, Yan W-W, Chan K, Chan W-M, Ngai C-W, Law K-I, Chow F-L, Liu R, Lai K-Y, Lau CCY, Liu S-H, Chan K-H, Lin C-K, Yuen K-Y. Hyperimmune IV immunoglobulin treatment: a multicenter double-blind randomized controlled trial for patients with severe 2009 influenza A(H1N1) infection. Chest 2013; 144: 464–473.

18. Group IFIPS. INSIGHT FLU005: An Anti-Influenza Virus Hyperimmune Intravenous Immunoglobulin Pilot Study. J Infect Dis 2016; 213: 574–578.

19. Beigel JH, Tebas P, Elie-Turenne M-C, Bajwa E, Bell TE, Cairns CB, Shoham S, Deville JG, Feucht E, Feinberg J, Luke T, Raviprakash K, Danko J, O’Neil D, Metcalf JA, King K, Burgess TH, Aga E, Lane HC, Hughes MD, Davey RT, Team IRCS. Immune plasma for the treatment of severe influenza: an open-label, multicentre, phase 2 randomised study. Lancet Respir Med 2017; 5: 500–511.

20. Delaney M, Wendel S, Bercovitz RS, Cid J, Cohn C, Dunbar NM, Apelseth TO, Popovsky M, Stanworth SJ, Tinmouth A, Van De Watering L, Waters JH, Yazer M, Ziman A, Biomedical Excellence for Safer Transfusion C. Transfusion reactions: prevention, diagnosis, and treatment. Lancet 2016; 388: 2825–2836.

21. Panch SR, Montemayor-Garcia C, Klein HG. Hemolytic Transfusion Reactions. The New England journal of medicine 2019; 381: 150–162.

